# Development of the Susceptibility-Spectrum Discrepancy Index (S2DI): A novel metric for antimicrobial stewardship in hospitalised patients

**DOI:** 10.64898/2026.03.23.26349044

**Authors:** Shinya Tsuzuki, Ryuji Koizumi, Yusuke Asai, Yuuki Hashimoto, Norihiko Inoue, Norio Ohmagari

## Abstract

**Objectives:** Optimising parenteral antimicrobial use is central to antimicrobial resistance (AMR) control, yet its appropriateness is difficult to assess. We aimed to develop a quantitative indicator to evaluate the appropriateness of parenteral antimicrobial therapy in hospitalised patients with bloodstream infections.

**Methods:** We developed the Susceptibility–Spectrum Discrepancy Index (S2DI), reflecting the discrepancy between antimicrobial susceptibility of blood culture isolates and the spectrum width of prescribed agents. Using a database from 67 National Hospital Organization hospitals in Japan, we identified patients with *Staphylococcus aureus* or *Escherichia coli* bacteraemia from 2017 to 2023. An expert panel of 10 infectious disease physicians independently ranked antimicrobial susceptibility (A) and spectrum width of commonly used agents (B). S2DI was defined as B minus A on day 7 after treatment initiation, with values closer to zero indicating more appropriate therapy. S2DI was calculated for individual cases, aggregated at the hospital level, and analysed using linear mixed-effects models with hospital-level random effects.

**Results:** A total of 4,505 *S. aureus* and 9,563 E. coli bacteraemia cases were included. Median S2DI was 1 (IQR 0–1) for *S. aureus* and 2 (IQR 0–3) for *E. coli.* For both pathogens, later calendar years were significantly associated with more favourable S2DI, suggesting gradual improvement in antimicrobial use. In *E. coli* bacteraemia, female sex and younger age were also associated with more appropriate therapy.

**Conclusions:** Although variation across hospitals persists, appropriateness of parenteral antimicrobial use has improved over time. S2DI is a simple metric that may support optimisation of antimicrobial use.

**Funding:** This study was supported by research grants from the Ministry of Health, Labour and Welfare, Japan (JP23HA1004 and JP22HA1004) and JSPS KAKENHI (JP23K18396).

## Introduction

In recent years, antimicrobial resistance (AMR) has been regarded as a global health threat [1,2]. Various measures have been implemented to reduce the disease burden caused by AMR, and the appropriate use of antimicrobial agents is one of the most fundamental [3,4].

In ambulatory care settings, antibiotics are predominantly prescribed in oral form and the most common cause of unnecessary antibiotic prescription is upper respiratory infections [5]. Most upper respiratory infections are viral, self-limiting diseases and prescribing antibiotics of any class to patients diagnosed solely with upper respiratory infections can be regarded as inappropriate. The appropriateness of antimicrobial use in ambulatory care settings can be evaluated retrospectively.

The evaluation of parenteral antimicrobials is more complex. Hospitalised patients are typically diagnosed with multiple conditions and receive ongoing treatment within the hospital setting. Consequently, the appropriateness of antimicrobial use during hospitalisation is often more difficult to assess.

The practice of ‘de-escalation’ further complicates the situation. When a critical infection such as bacteraemia is suspected, physicians initiate empiric antibiotic therapy alongside submitting culture tests, switching to definitive therapy once the culture results are available. This approach is believed to curb unnecessary use of broad-spectrum antibiotics and prevent the spread of AMR [6,7]. In settings in which such practices are common, the appropriateness of antimicrobial use becomes difficult to assess.

As previously stated, promoting antimicrobial stewardship is considered to be directly linked to AMR countermeasures. However, the appropriateness of the use of parenteral antimicrobials at the point when definitive therapy becomes available has scarcely been evaluated at the population level to date. The objective of this study was to establish indicators for evaluating the appropriateness of parenteral antimicrobial use at the facility level, enabling comparisons across multiple facilities and at the national level.

## Methods

### Data source and settings

In this retrospective cohort study, we developed the Susceptibility–Spectrum Discrepancy Index (S2DI), a novel metric that captures the discrepancy between antibiotic susceptibility of the organisms detected by blood culture and the width of the spectrum of the antimicrobials prescribed. Details of the scoring system for the S2DI are discussed later.

We used a database comprising 67 National Hospital Organization hospitals in Japan and extracted patients for whom *S. aureus* or *E. coli* was detected in blood cultures between 2017 and 2023. Patients from whom multiple bacteria were detected were excluded. As described later, our primary interest was the appropriateness of treatment 7 days after the detection of bacteria in blood cultures. Therefore, we also excluded cases that were discharged or died within 7 days from the blood culture collection. We aggregated the characteristics of facilities (the number of beds and location) and blood-culture positive patients (age, sex, Charlson comorbidity index [CCI], date of admission, date of blood culture submission, date of bacteria detection in blood specimen, start date of antibiotic therapy, antibiotics used after 7 days from the date of blood culture collection, and their clinical outcomes [e.g., discharged alive, transferred, etc.]). We used the date of blood culture collection as the proxy of the day of bacterial detection because the database did not include this information.

### Scoring methodology of the S2DI

We ranked the antimicrobial susceptibility for each bacterium (A) and the width of the spectrum of antimicrobials (B) through an expert panel of 10 infectious disease physicians from the Disease Control and Prevention Center, National Center for Global Health and Medicine, Japan Institute for Health Security, which is located in Tokyo and one of six national centre hospitals in Japan. The antibiotic susceptibility of *S. aureus* is classified into three levels, with corresponding antimicrobials ranked into four levels; whereas the antibiotic susceptibility of *E. coli* is classified into five levels, with corresponding antimicrobials ranked into six levels. The details of the ranking are available in the appendix (Supplementary file 1).

Notably, the S2DI score was not designed to evaluate the appropriateness of empiric therapy immediately after the diagnosis of bacteraemia. This scoring system was devised solely to assess the appropriateness of treatment when de-escalation has been conducted, and the susceptibility of the detected bacterium has been determined. Furthermore, to prevent diagnostic uncertainty, this study restricted its subjects to cases of confirmed bacteraemia caused by *E. coli* or *S. aureus*, i.e., bacteria not considered contaminants in blood culture.

### Statistical analysis

The characteristics of the facilities included in the dataset and those of the extracted patients are presented as the median and interquartile range (IQR). In addition, we exploratively identified factors correlated with the S2DI score using a liner mixed-effects model accounting for differences in each facility as random effects. We included calendar year, number of beds (as a categorical variable divided into two categories: under 400 beds and 400 beds or more), location of facilities (regional level), whether or not the patient was being treated by a specific medical department (infectious diseases, paediatrics, or psychiatry), and the patients’ sex, age, and CCI [8,9] as variables for the fixed effect.

All statistical analyses were conducted using R, version 4.2.2 [10]. A two-sided *p* value lower than 0.05 was considered statistically significant.

### Ethics

This study was approved by the Certified Review Board of Japan Institute for Health Security (Approval number: JIHS-S-004611-04). The study was conducted in accordance with the Declaration of Helsinki.

## Results

A total of 4,505 *S. aureus* and 9,563 *E. coli* bacteraemia cases were included. Table 1-a and 1-b shows the characteristics of facilities included because of bacteraemia cases caused by *E. coli* and *S. aureus*. The data for all cases extracted from the database (i.e., cases that were discharged or died within 7 days from the day of blood culture collection were excluded) are available in Supplementary file 2. Among the 67 hospitals included in the database, approximately (varied by year) 50 facilities included bacteraemia cases caused by *E. coli* and/or *S. aureus*. Most facilities had dedicated departments for paediatrics and psychiatry, while only 10 or 11 facilities had a department for infectious diseases. The data were collected from all regions of Japan. For reference, a regional map of Japan is provided as Figure 1.

**Figure 1.**
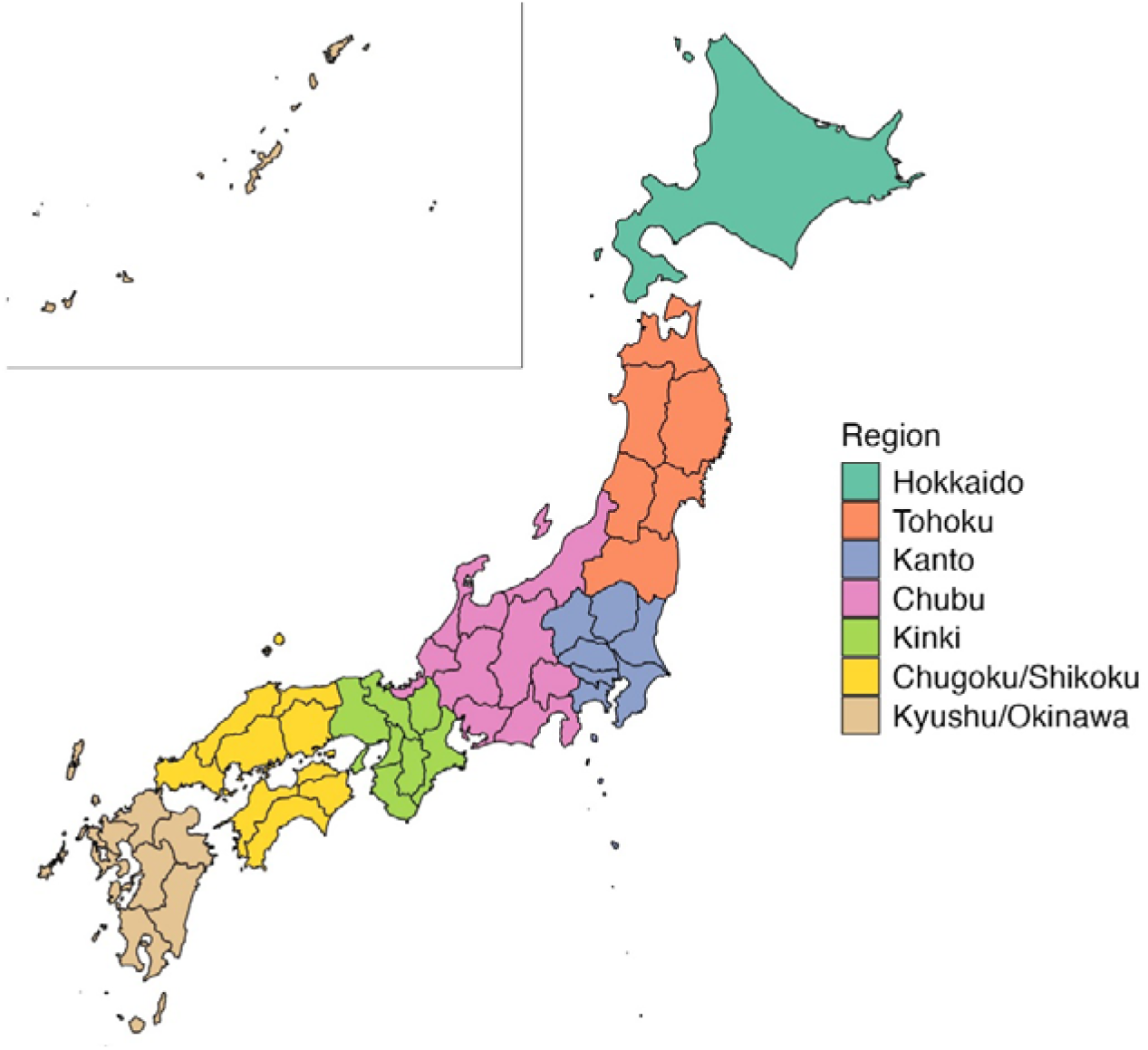
Geographic classification of participating facilities across Japan. Japan was divided into seven standard geographic regions—Hokkaido, Tohoku, Kanto, Chubu, Kinki, Chugoku/Shikoku, and Kyushu/Okinawa—based on administrative prefectures. Colours indicate the region where each facility is located. This classification was used for regional comparisons in the mixed-effects models.

**Table 1-a.** Characteristics of the facilities from which the bacteraemia cases caused by *Escherichia coli* originated.

| **Year** | **2017** | **2018** | **2019** | **2020** | **2021** | **2022** | **2023** | **Total** |
| --- | --- | --- | --- | --- | --- | --- | --- | --- |
| **Number of facilities** | 44 | 46 | 51 | 54 | 57 | 53 | 58 | 61 |
| **Department of infectious disease** | 10 | 10 | 10 | 11 | 11 | 11 | 11 | 11 |
| **Department of paediatrics** | 41 | 43 | 47 | 49 | 52 | 48 | 53 | 56 |
| **Department of psychiatry** | 34 | 36 | 39 | 39 | 41 | 39 | 42 | 43 |
| **Hokkaido region** | 3 | 4 | 4 | 4 | 4 | 4 | 4 | 4 |
| **Tohoku region** | 3 | 3 | 3 | 3 | 4 | 2 | 4 | 4 |
| **Kanto region** | 8 | 9 | 8 | 11 | 11 | 10 | 11 | 13 |
| **Chubu region** | 8 | 7 | 8 | 8 | 8 | 7 | 8 | 8 |
| **Chugoku/Shikoku region** | 8 | 8 | 10 | 10 | 10 | 11 | 11 | 12 |
| **Kinki region** | 4 | 5 | 6 | 6 | 8 | 7 | 8 | 8 |
| **Kyushu/Okinawa region** | 10 | 10 | 12 | 12 | 12 | 12 | 12 | 12 |
| **<400 beds** | 16 | 16 | 20 | 23 | 23 | 20 | 24 | 26 |
| **≥400 beds** | 28 | 30 | 31 | 31 | 34 | 33 | 34 | 35 |

**Table 1-b.** Characteristics of the facilities from which the bacteraemia cases caused by *Staphylococcus aureus* originated.

| **Year** | **2017** | **2018** | **2019** | **2020** | **2021** | **2022** | **2023** | **Total** |
| --- | --- | --- | --- | --- | --- | --- | --- | --- |
| **Number of facilities** | 45 | 47 | 50 | 51 | 53 | 53 | 56 | 61 |
| **Department of infectious disease** | 10 | 10 | 10 | 11 | 11 | 11 | 11 | 11 |
| **Department of paediatrics** | 42 | 44 | 45 | 48 | 48 | 49 | 52 | 56 |
| **Department of psychiatry** | 35 | 38 | 38 | 39 | 39 | 40 | 39 | 43 |
| **Hokkaido region** | 3 | 4 | 4 | 4 | 4 | 4 | 4 | 4 |
| **Tohoku region** | 3 | 2 | 3 | 2 | 2 | 2 | 4 | 4 |
| **Kanto region** | 11 | 11 | 9 | 13 | 12 | 13 | 13 | 13 |
| **Chubu region** | 7 | 8 | 8 | 7 | 8 | 8 | 7 | 8 |
| **Chugoku/Shikoku region** | 6 | 7 | 9 | 7 | 10 | 8 | 11 | 12 |
| **Kinki region** | 5 | 5 | 6 | 6 | 5 | 6 | 5 | 8 |
| **Kyushu/Okinawa region** | 10 | 10 | 11 | 12 | 12 | 12 | 12 | 12 |
| **<400 beds** | 16 | 17 | 18 | 19 | 23 | 22 | 22 | 26 |
| **≥400 beds** | 29 | 30 | 32 | 32 | 30 | 31 | 34 | 35 |

Table 2-a and 2-b shows the characteristics of the *E. coli* and *S. aureus* bacteraemia patients extracted from the database. The median age of *E. coli* bacteraemia cases was 80 (interquartile range [IQR] 72–86), while that of *S. aureus* bacteraemia cases was 76 (64–84). The percentage of female patients was greater for *E. coli* bacteraemia cases than that for *S. aureus* bacteraemia cases (55.1% vs 37.1%, respectively). The median score for the CCI in both groups was 1. The 30-day all-cause mortality rate in *E. coli* bacteraemia patients was 3.3%, while that in *S. aureus* bacteraemia patients was 9.5%. The median number of days from admission to the detection of bacteria was 1 (IQR 1–2) day and 3 (IQR 1–17) days for the *E. coli* and *S. aureus* groups, respectively. The total length of hospital stay of *E. coli* bacteraemia patients was 19 (IQR 13–34) days, while that of *S. aureus* bacteraemia patients was 42 (IQR 24–69) days. The median score of the S2DI in *E. coli* bacteraemia cases was 2 (IQR 0–3), while that in *S. aureus* bacteraemia cases was 1 (IQR 0–1).

**Table 2-a.**
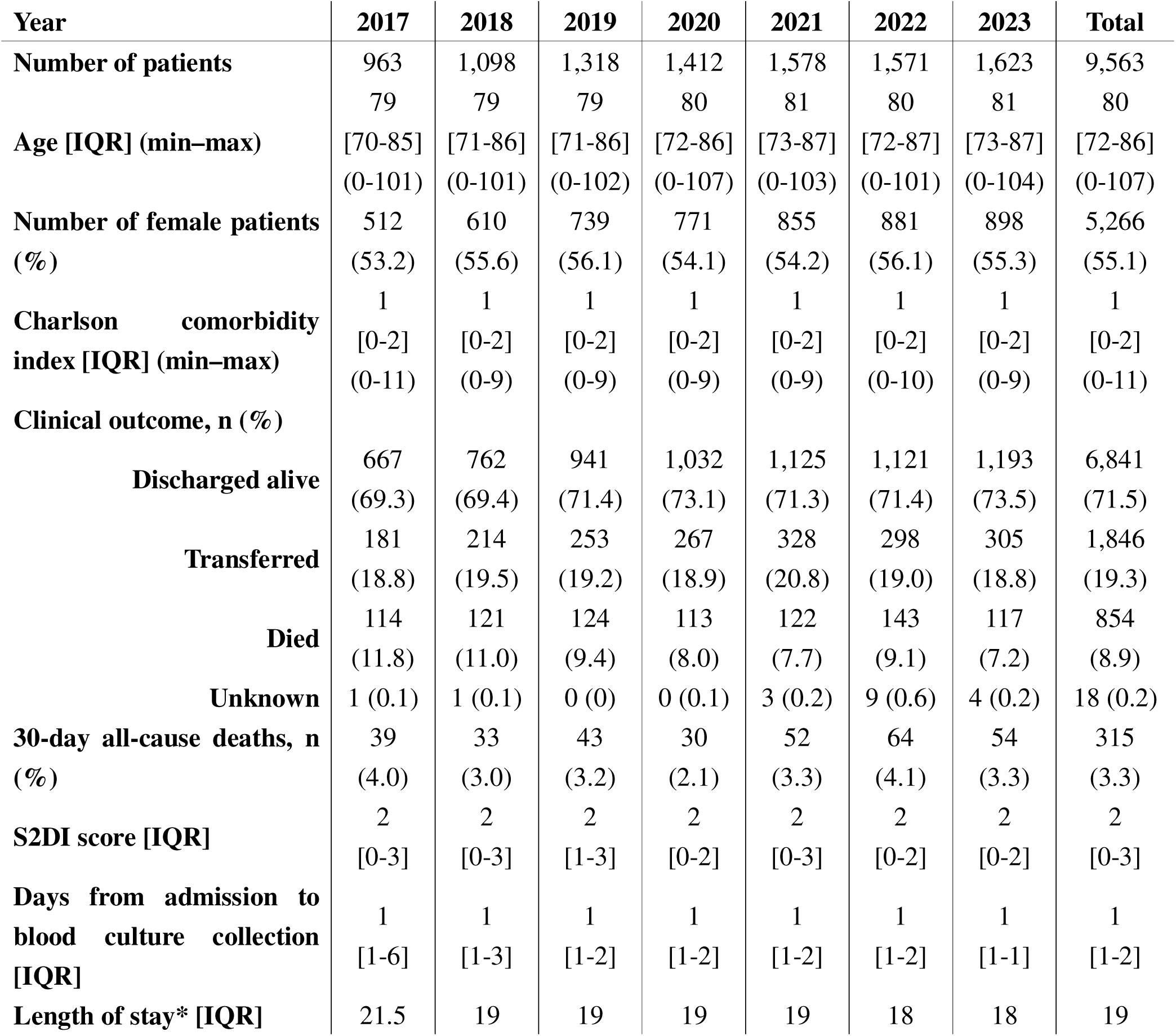
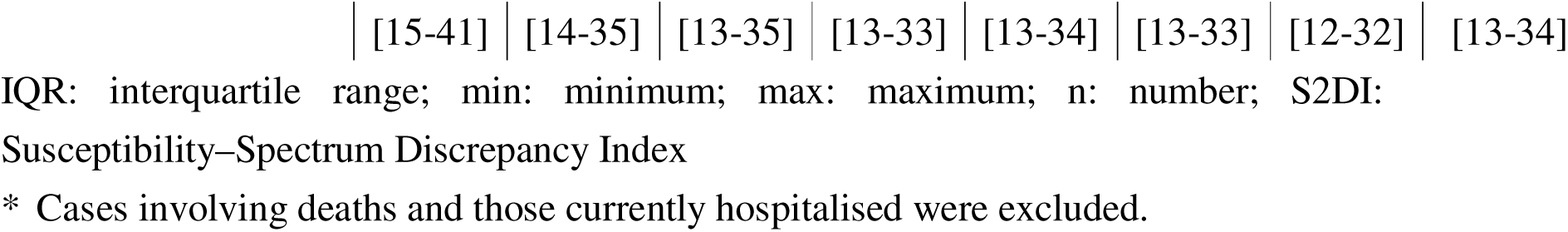
Characteristics of the *Escherichia coli* bacteraemia patients extracted from the database.

**Table 2-b.**
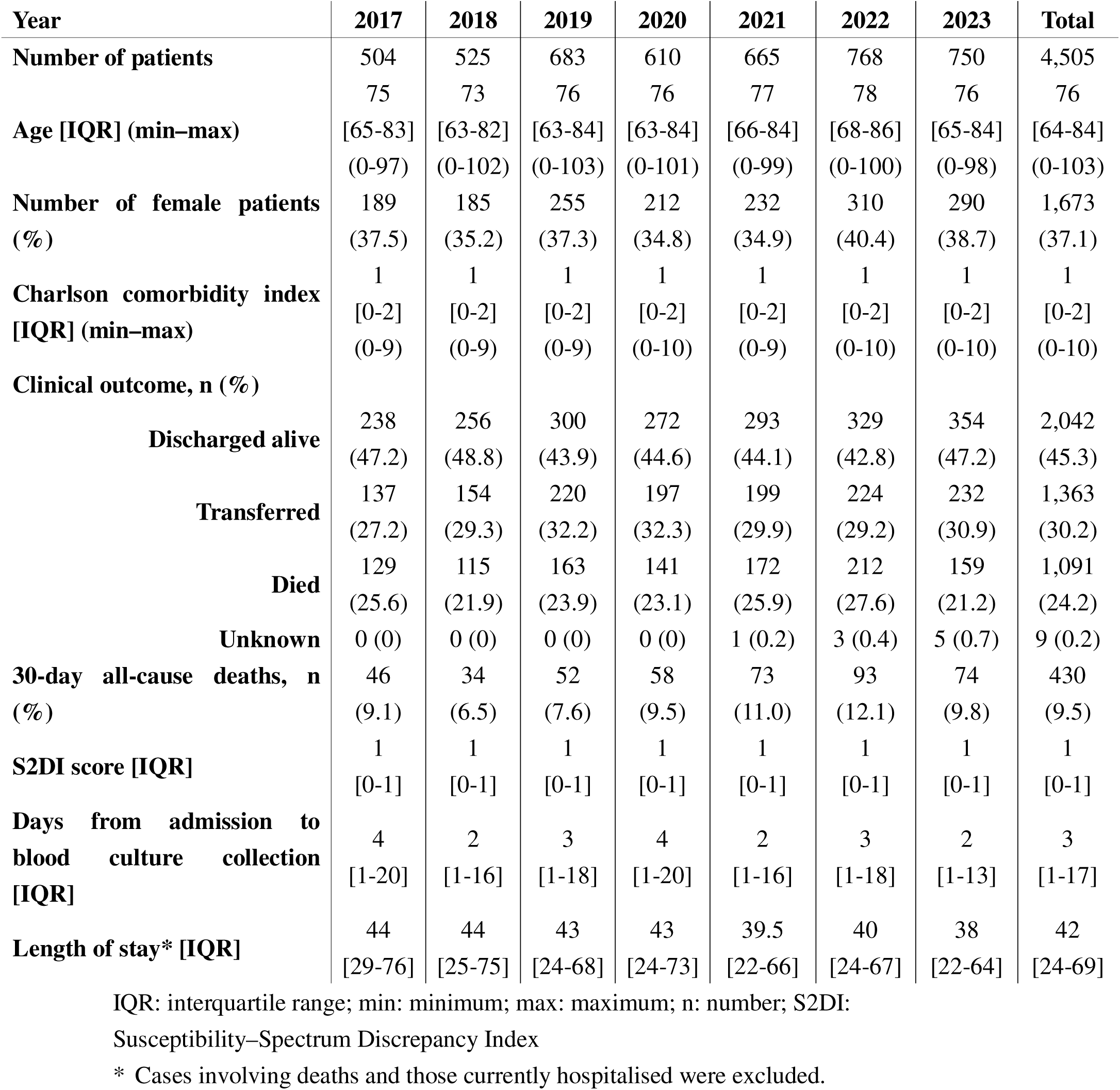
Characteristics of the *Staphylococcus aureus* bacteraemia patients extracted from the database.

Figure 2 demonstrates the daily S2DI scores from the initiation of the treatment for bacteraemia. Disposition outcomes including discharge, transfer, and death are also available in Supplementary file 2.

**Figure 2.**
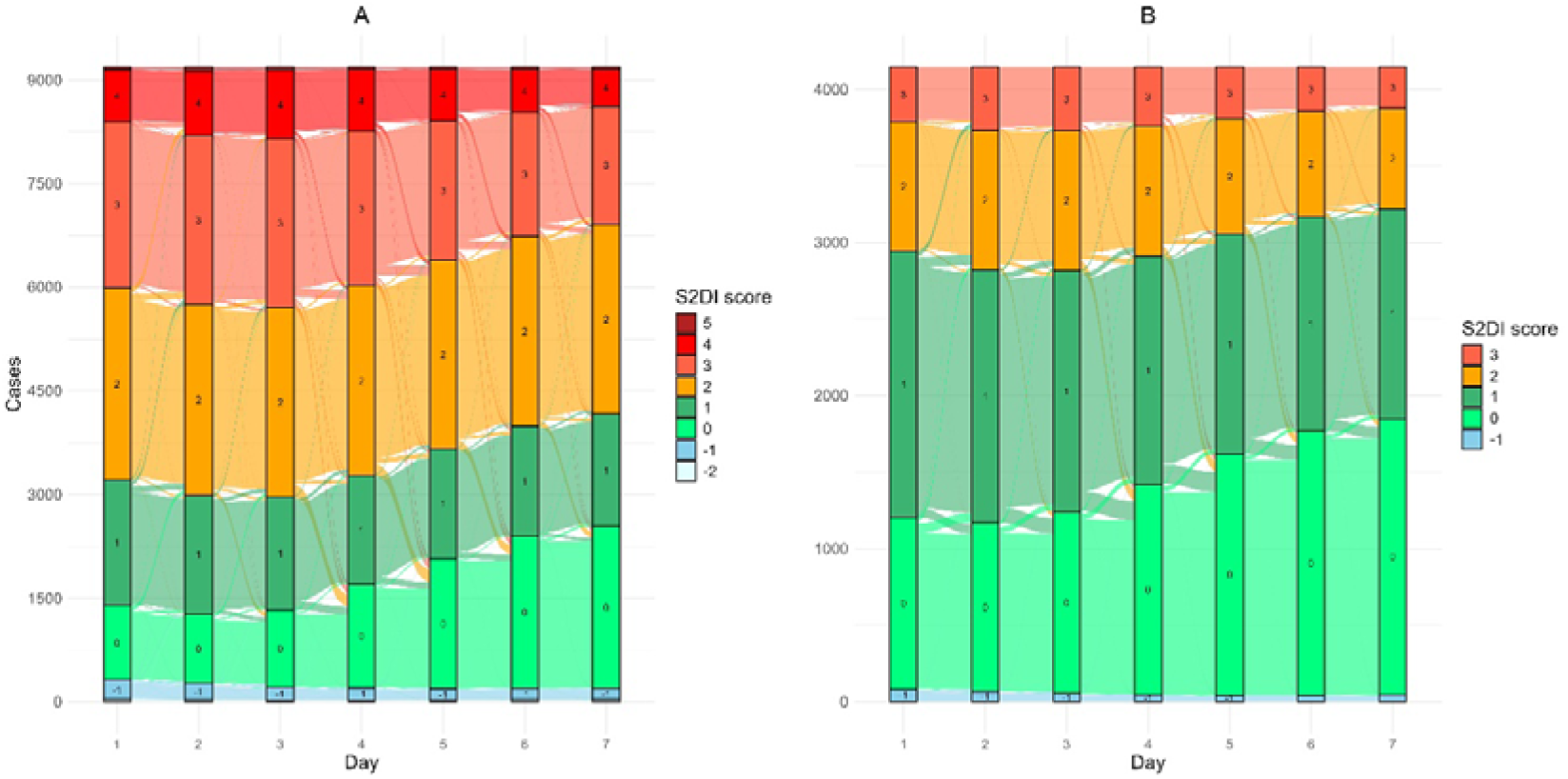
Daily transitions of the Susceptibility–Spectrum Discrepancy Index (S2DI) scores during the first 14 days from the treatment initiation of *Escherichia coli* (left panel) and *Staphylococcus aureus* (right panel) bacteraemia. Stacked bar charts show the distribution of S2DI scores on each hospital day. Coloured ribbons illustrate the day-to-day transitions in S2DI scores for individual patients.

Table 3-a and 3-b shows the characteristics of the patients by S2DI score.

**Table 3-a.**
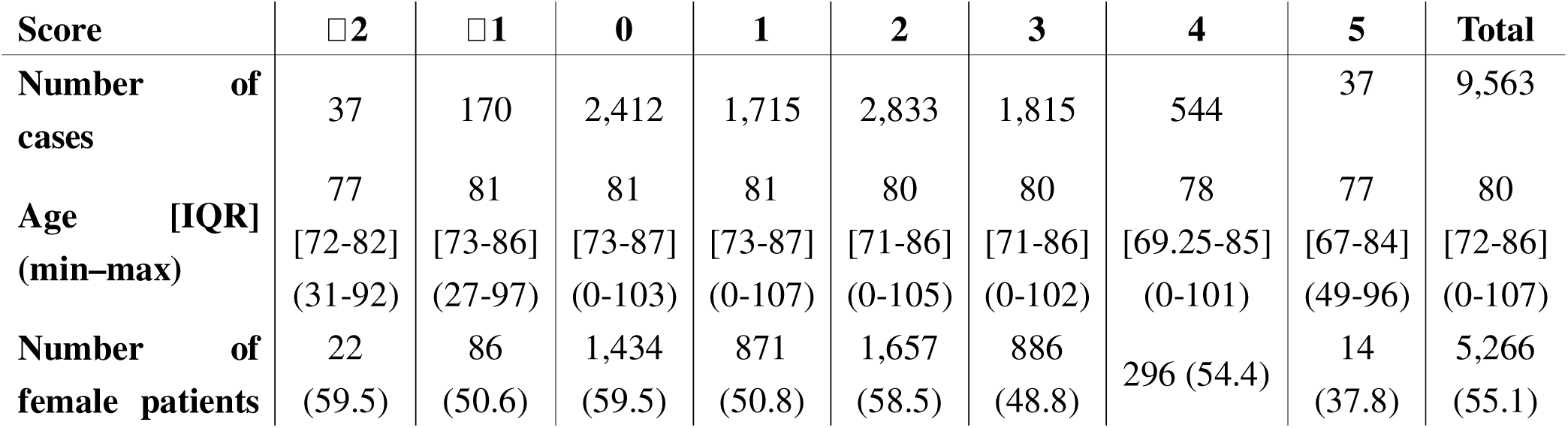
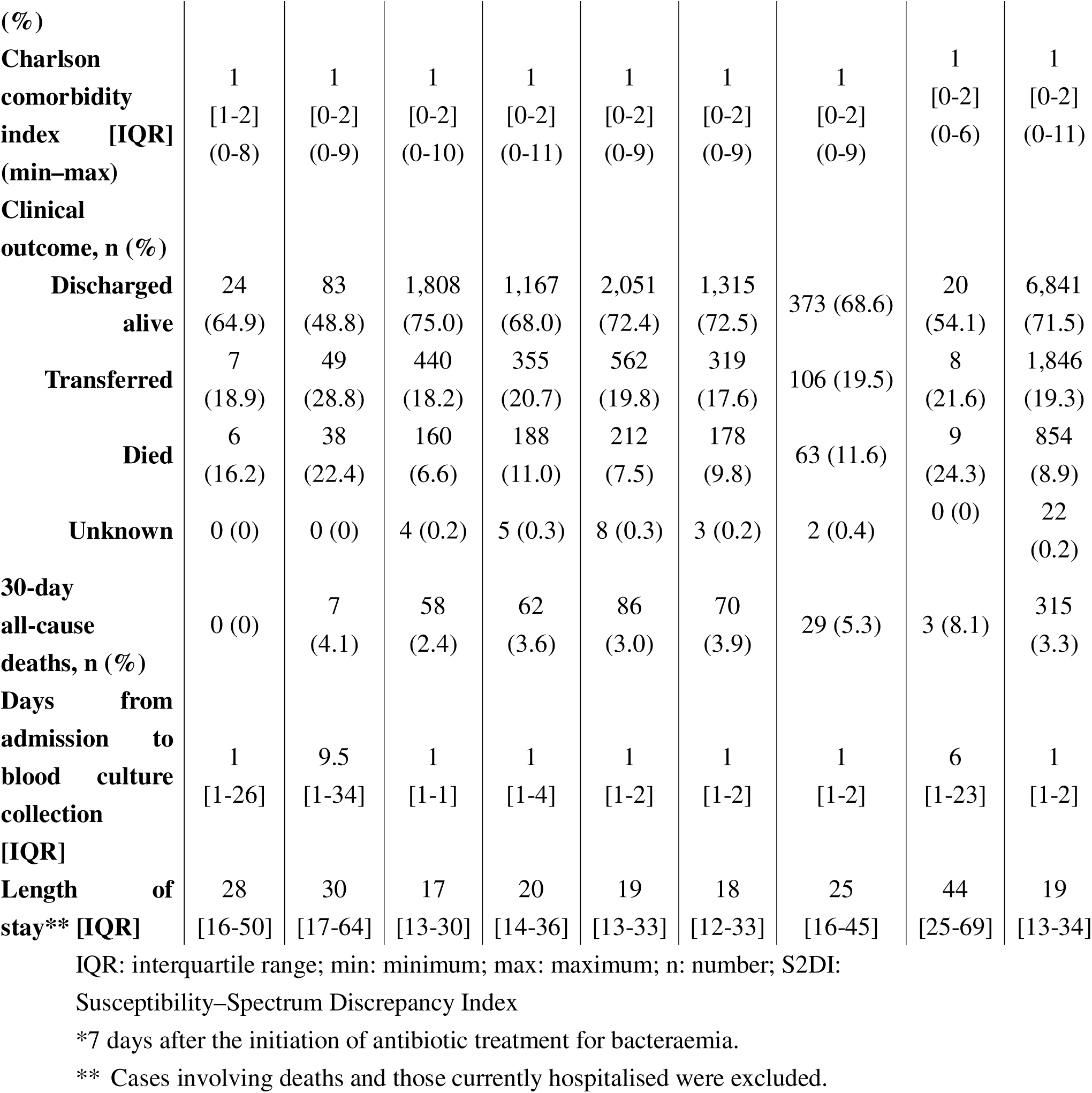
Characteristics of the *Escherichia coli* bacteraemia patients by S2DI score at Day 7*.

**Table 3-b.**
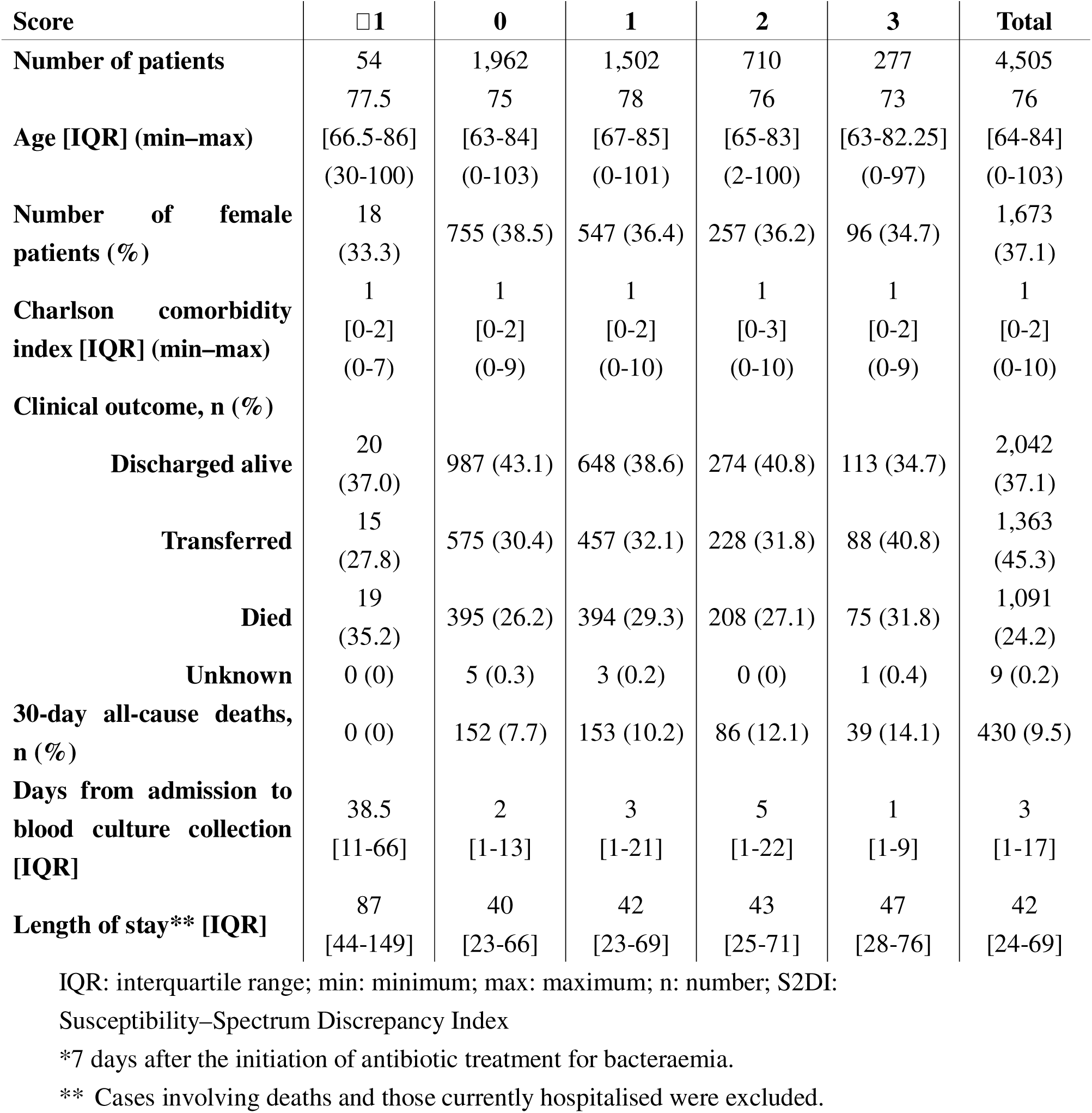
Characteristics of the *Staphylococcus aureus* bacteraemia patients by S2DI score at Day 7*.

The linear-mixed effects model suggested that the number of cases of both *S. aureus* and *E. coli* bacteraemia per calendar year were increasing annually, along with a favourable S2DI score, which exhibited a tendency for improved scores in later years (*p* < 0.001 for *E. coli* bacteraemia, *p* < 0.002 for *S. aureus* bacteraemia). For *E. coli*, in addition to calendar year, female sex (*p* < 0.001) and younger age (*p* < 0.001) also correlated with favourable scores, whereas a facility having a department of infectious disease was associated with favourable scores for *S. aureus* bacteraemia (*p* = 0.008). The estimated correlations between S2DI score and factors included in the model as fixed effects are presented in Figure 3.

**Figure 3.**
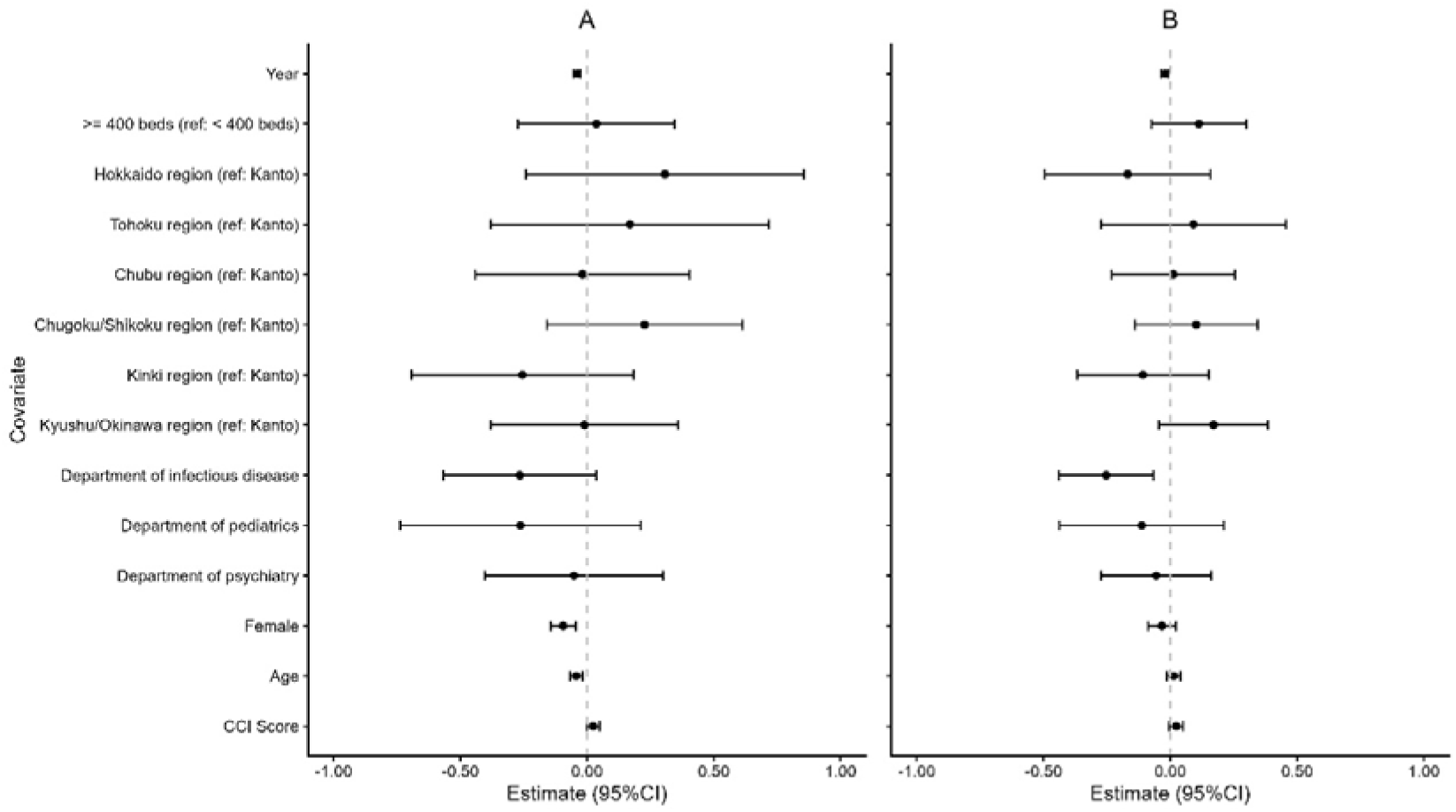
Forest plots of fixed-effect estimates from linear mixed-effects models for *Escherichia coli* (left panel) and *Staphylococcus aureus* (right panel) bacteraemia. Each point represents the adjusted fixed-effect estimated coefficients, with horizontal bars showing the corresponding 95% confidence intervals. Covariates include patient-level factors (age, sex, Charlson comorbidity index), specific departments at facilities, hospital bed size, geographic region (reference: Kanto, in which Tokyo is located), and calendar year. Positive estimates indicate a relative tendency toward broad-spectrum antibiotic use, whereas negative estimates indicate a tendency toward narrow-spectrum therapy. Estimates crossing zero suggest no statistically significant association.

To assess between-facility variability in antibiotic prescribing behaviour, we constructed caterpillar plots (Figure 4) based on the best linear unbiased predictions (BLUPs) of the facility-level random intercepts from the models. Random intercepts represent facility-specific deviations from the overall mean on the log-odds scale. For each facility, BLUPs and their approximate 95% confidence intervals were plotted and sorted by magnitude. Positive BLUPs denote a stronger inclination toward broad-spectrum therapy, whereas negative BLUPs indicate a relative preference for narrow-spectrum regimens. A random intercept among hospitals showed a variance of 0.168 (standard deviation [SD] 0.409) for *E. coli* bacteraemia and a variance of 0.053 (SD 0.230) for *S. aureus* bacteraemia. The intra-cluster correlation for *E. coli* bacteraemia and *S. aureus* bacteraemia was 0.104 and 0.062, respectively.

**Figure 4.**
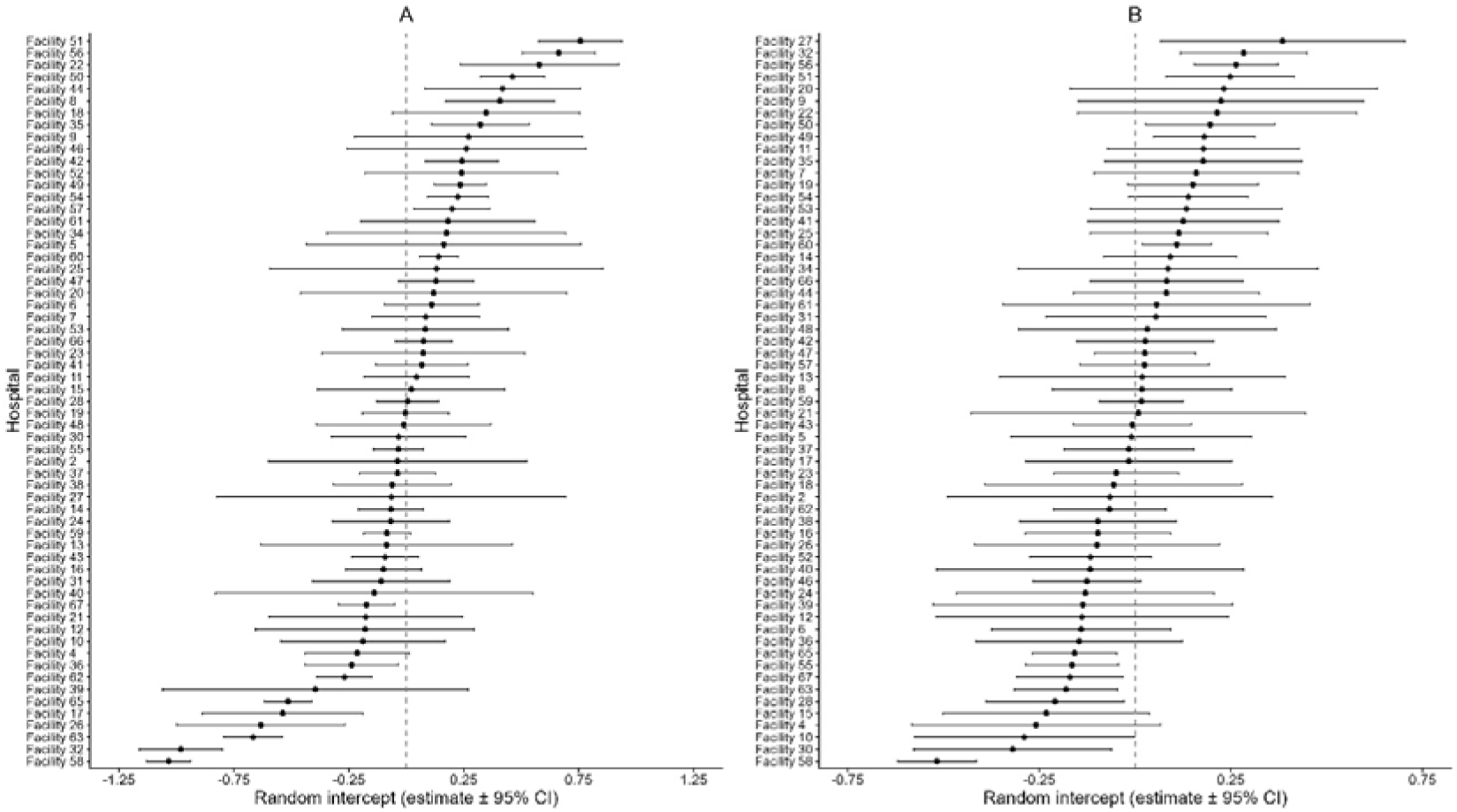
Caterpillar plots of facility-level random intercepts from the mixed-effects models for *Escherichia coli* (left panel) and *Staphylococcus aureus* (right panel) bacteraemia. Each point represents the best linear unbiased prediction (BLUP) of the facility-specific random intercept, with horizontal bars indicating the corresponding 95% confidence intervals. The random intercepts are centred at zero, which represents the average facility. Positive values indicate a greater tendency toward broad-spectrum antibiotic use, whereas negative values indicate a tendency toward narrow-spectrum use. Facilities are ordered by the magnitude of their BLUPs.

To illustrate between-facility variability in antibiotic prescribing tendencies, we generated snail plots based on BLUPs of the facility-level random intercepts from the mixed-effects models (Figure 5). Snail plots display the BLUPs in a radial layout in which the direction (positive vs negative) indicates a relative tendency toward broad– or narrow-spectrum antibiotic use, and the radial length represents the magnitude of the random effect.

**Figure 5.**
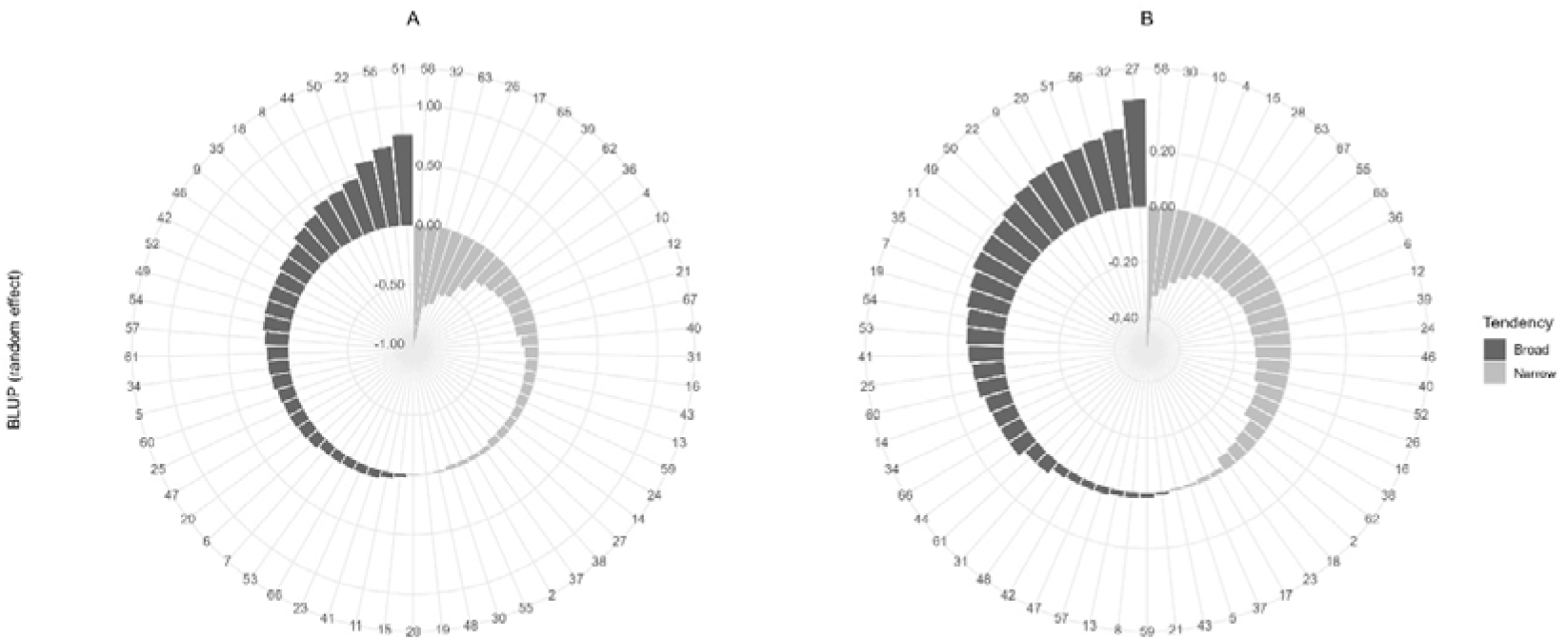
Snail plots of facility-level random effects for broad– and narrow-spectrum antibiotic use in cases of *Escherichia coli* (left panel) and *Staphylococcus aureus* (right panel) bacteraemia. Each bar represents the best linear unbiased prediction (BLUP) of the facility-level random intercept from the mixed-effects model. Bars extending upward indicate a tendency toward broad-spectrum antibiotic use, whereas downward bars indicate a tendency toward narrow-spectrum antimicrobials. The magnitude of each bar reflects the relative contribution of each facility to between-facility variability. Dark bars denote broad-spectrum-leaning facilities and light bars denote narrow-spectrum-leaning facilities. Facility identifiers are shown around the circumference.

### Sensitivity analysis

We conducted sensitivity analyses with the same model used for the main results, using data exclusively from facilities that provided all data for the 7-year period from 2017 to 2023 to the database. The results showed no significant difference from the main analysis, except that no significant association was found between a favourable S2DI score for *S. aureus* bacteraemia and the facility having a department of infectious disease (*p* = 0.104). The details of the results are available in Supplementary file 2.

## Discussion

To our knowledge, few previous studies have examined indicators that enable the assessment of the appropriateness of parenteral antibiotic use at the facility level and facilitate comparison across multiple facilities. This is likely attributable, as previously stated, to the difficulty of assessing the appropriateness of antimicrobial use in individual hospitalised cases.

Cicollini *et al*. used national surveillance data to create a ‘Drug Effectiveness Index’ based on the prevalence of microorganisms, resistance rates, and the frequency of antimicrobial use, thereby evaluating appropriateness at the population level [11]. This approach is simpler than evaluating individual cases; however, because it does not serve as an indicator for comparing the appropriateness of antimicrobial use at the facility level, it cannot identify which facilities within the country are excelling in appropriate use or which have room for improvement.

Conversely, S2DI evaluates the appropriateness of antimicrobial use in individual cases and aggregates these data by facility. This enables the assessment of the relative strengths of each facility’s antimicrobial stewardship programme. In this respect, S2DI can be considered superior as an indicator for AMR countermeasures. Although van den Bosch *et al*. also examined the applicability of generic quality indicators at the facility level [12], their studies assessed the applicability of existing evaluation metrics [13] and did not directly evaluate the appropriateness of antimicrobial use at individual facilities.

The descriptive S2DI score aggregation revealed that even 7 days after the initiation of bacteraemia treatment, numerous cases had not undergone optimisation of empirical therapy, and this applied to every facility. This suggests that the appropriate use of parenteral antimicrobials in Japan still has room for improvement.

However, the results of multivariable analysis using mixed-effects models revealed a tendency for S2DI scores to improve over time—an insight that would have been difficult to detect from simple aggregation alone. This validates our focus on AMR countermeasures since publication of the National Action Plan [14].

The results of the linear mixed-effects models also suggested that the appropriate use of parenteral antimicrobials varies significantly between facilities, and evaluating each facility individually can identify those requiring priority intervention. Although this study does not aim to explicitly identify facilities where antimicrobial stewardship is lagging, prioritising intervention in facilities with poor S2DI scores could conceivably advance appropriate use at the national level.

The mixed-effects models also indicated that female sex, younger age, and lower CCI scores correlated with favourable S2DI scores in *E. coli* bacteraemia, potentially reflecting the perception of clinicians that de-escalation is more easily achieved in younger patients with fewer underlying conditions. Nevertheless, because these findings were not observed in *S. aureus* patients, we should be cautious in interpreting these results. Caution should also be exercised regarding the correlation between facilities having a department of infectious disease and favourable S2DI scores in patients with *S. aureus* bacteraemia.

The absence of any association between low S2DI scores and increased 30-day and all-cause mortality rates may also be considered a positive finding. Reports from several countries, including Japan, indicate that de-escalation does not worsen patient outcomes [7,15,16]. Our findings may serve as a renewed justification for clinicians to actively pursue de-escalation. However, it should be noted that this result is merely incidental and has not been adjusted for important factors such as disease severity.

Our study has several limitations. First, the S2DI scoring system is intended solely for bacteraemia caused by *E. coli* and *S. aureus*, and does not cover other common diseases such as pneumonia caused by *Streptococcus pneumoniae*. To assess the appropriateness of parenteral antibiotic use for other diseases, we would need to develop other rankings for both antibiotics and susceptibility. However, as previous studies demonstrated, bacteraemia, particularly that caused by *E. coli* and *S. aureus*, accounts for the majority of the disease burden attributable to AMR [2,17,18], a phenomenon that is especially eminent in Japan [19–21]. Therefore, there is considerable justification for prioritising the assessment of the appropriateness of antimicrobial use for bacteraemia caused by these two bacteria.

Second, this study is retrospective using health record data. However, the nature of the data made it difficult to accurately assess disease severity. Factors that may have affected disease severity, such as de-escalation and allergies and/or adverse effects due to a specific class of antibiotics, would not be accounted for by the S2DI score.

As with other studies employing mixed-effects models, unmeasured confounding factors are likely to exist [22] and should be considered when interpreting the results. Furthermore, the effect of the S2DI score might be non-linear because it is an ordinary scale indicator. Selection biases due to the elimination of fatal cases within 7 days from the day of admission may also influence the results. Nevertheless, the fact that we have quantitatively demonstrated disparities in antimicrobial stewardship between facilities is an interesting finding that may benefit future AMR countermeasures.

In conclusion, the S2DI provides a straightforward, quantitative measure that may assist clinicians and health policymakers in monitoring and enhancing antimicrobial stewardship, albeit with certain limitations to be considered. In this study, assessment by S2DI revealed significant inter-facility variation in antimicrobial stewardship, whilst also suggesting that appropriate antimicrobial use may be increasing annually.

## Author contributions

ST conceived the study. RK, YH, and NI curated and analysed the data. ST and YA validated the results. ST, RK, and YA interpreted the results. ST wrote the first draft of the manuscript and other authors critically reviewed and revised it. NO supervised the research project. All authors have read and agreed to the final version of the manuscript.

## Transparency declaration

The authors declare that they have no conflicts of interest. This study was supported by research grants from the Ministry of Health, Labour and Welfare, Japan (JP23HA1004 and JP22HA1004) and JSPS KAKENHI (JP23K18396).

## Data sharing

The datasets generated and analysed for this study are not publicly available due to the inclusion of sensitive personal information. The analytic code is available from the corresponding author upon reasonable request.

## Acknowledgements

We would like to express our sincere gratitude to Drs Kayoko HAYAKAWA, Shinichiro MORIOKA, Yutaro AKIYAMA, Masahiro ISHIKANE, Noriko IWAMOTO, Yuki MORIYAMA, Hidetoshi NOMOTO, Sho SAITO, Aki SAKURAI, and Kei YAMAMOTO (Disease Control and Prevention Center, National Center for Global Health and Medicine Hospital, Japan Institute for Health Security) for their contributions as members of the S2DI scoring system expert panel. We thank Edanz (https://jp.edanz.com/ac) for editing a draft of this manuscript.

