## Supplementary File 1 for "Development of the Susceptibility-Spectrum Discrepancy Index (S2DI): A novel metric for antimicrobial stewardship in hospitalised patients"

**Supplementary information for**

**Development of the Susceptibility–Spectrum Discrepancy Index (S2DI): A novel metric for antimicrobial stewardship in hospitalised patients**

Shinya Tsuzuki, Ryuji Koizumi, Yusuke Asai, Yuuki Hashimoto, Norihiko Inoue, Yusuke Asai, Norio Ohmagari

The definition of the Susceptibility–Spectrum Discrepancy Index (S2DI) score:

(A) rank of the antimicrobial susceptibility of bacteria (higher rank indicates higher resistance)

(B) rank of the spectrum of antimicrobials (higher rank indicates a broader spectrum)

S2DI score = (B) ˗ (A)

A score closer to zero indicates more appropriate treatment, while a higher score signifies the unnecessary use of broader-spectrum antibiotics.

The S2DI scoring system expert panel, which consisted of 10 infectious disease physicians (named in the Acknowledgments section in the main text) from the Disease Control and Prevention Center at the National Center for Global Health and Medicine Hospital (Japan Institute for Health Security, Tokyo, Japan), discussed the rank of antibiotics and the characteristics of the bacteria for i) *Staphylococcus aureus* bacteraemia and ii) *Escherichia coli* bacteraemia cases.

Each panel member ranked the antibiotics and bacterial susceptibility, with the corresponding author collating the results. For items where opinions diverged, discussions were held until a consensus was reached, with reference to the mean and median values, and the authors determined the final ranking.

The ranking varies for each microorganism, and even for the same drug. For example, the rank of a drug as a treatment for *S. aureus* bacteraemia is not necessarily the same as its rank as a treatment for *E. coli* bacteraemia.

Rank of *Staphylococcus aureus*

**Rank 1:** methicillin-susceptible *S. aureus* (MSSA)

**Rank 2:** methicillin-resistant *S. aureus* (MRSA)

**Rank 3:** vancomycin-intermediate/resistant *S. aureus* (VISA/VRSA)

Rank of antibiotics for the treatment of *S. aureus* bacteraemia

**Rank 1:** benzylpenicillin, ampicillin, ampicillin and cloxacillin, cefazolin

**Rank 2:** azithromycin, amikacin, arbekacin, ampicillin/sulbactam, clindamycin, streptomycin, sulfamethoxazole and trimethoprim, cefotaxime, cefotiam, ceftriaxone, cefmetazole, tobramycin, vancomycin, piperacillin, flomoxef, fosfomycin, metronidazole

**Rank 3:** ciprofloxacin, cefepime, cefozopran, cefoperazone/sulbactam, ceftazidime, daptomycin, teicoplanin, pazufloxacin, minocycline, linezolid, levofloxacin

**Rank 4:** imipenem/cilastatin, piperacillin/tazobactam, doripenem, biapenem, meropenem

Rank of *Escherichia coli*

**Rank 1:** susceptible to ampicillin (ABPC) and/or cefazolin (CEZ)

**Rank 2:** not susceptible to ABPC and CEZ, but susceptible to ampicillin/sulbactam (ABPC/SB)

**Rank 3:** not susceptible to ABPC, CEZ, and ABPC/sulfamethoxazole and trimethoprim (ST), but susceptible to any of ceftriaxone (CTRX), ST, cefmetazole (CMZ), ceftazidime (CAZ), or amikacin (AMK)

**Rank 4:** not susceptible to ABPC, CEZ, ABPC/ST, CTRX, ST, CMZ, CAZ, and AMK, but susceptible to any of piperacillin/tazobactam (PIPC/TAZ), cefepime (CFPM), or levofloxacin (LVFX)

**Rank 5:** not susceptible to any of the drugs listed above, but susceptible to meropenem (MEPM)

Rank of antibiotics for the treatment of *E. coli* bacteraemia

**Rank 1:** benzylpenicillin, ABPC, CEZ

**Rank 2:** ABPC/ST, erythromycin, cefotiam, tobramycin

**Rank 3:** azithromycin, AMK, arbekacin, isepamicin, clindamycin, gentamicin, dibekacin, ST, cefotaxime, CAZ, CTRX, CMZ, PIPC, flomoxef, fosfomycin, metronidazole

**Rank 4:** ciprofloxacin, CFPM, cefozopran, cefoperazone/sulbactam, PIPC/TAZ, minocycline, LVFX

**Rank 5:** imipenem/cilastatin, ceftolozane/tazobactam, doripenem, biapenem, meropenem

**Rank 6:** daptomycin, linezolid, teicoplanin, vancomycin

The S2DI was defined as B minus A after 7 days from the day of bacteria detection in the blood specimen, with scores closer to zero indicating more appropriate antimicrobial use. For example, methicillin-susceptible *S. aureus* (MSSA) is classified into Rank 1 (A = 1). If a patient who had bacteraemia caused by MSSA is treated with vancomycin 7 days after MSSA was detected in the blood culture, B = 2 because vancomycin is classified into Rank 2. In this case, the S2DI score 2 ˗ 1 = 1 is assigned. By contrast, if the same patient is treated with cefazolin 7 days after MSSA detection, A = 1 because MSSA is Rank 1, and B = 1 because cefazolin is classified into Rank 1; therefore, the S2DI score 1 ˗ 1 = 0 is assigned. If another patient had methicillin-resistant *S. aureus* (MRSA) bacteraemia, A = 2 because MRSA is classified into Rank 2 and if this patient is treated with vancomycin, the S2DI score is also 0 (2 ˗ 2 = 0). Fundamentally, the rankings for both bacteria and antimicrobials are set such that the S2DI score becomes zero when appropriate treatment is administered according to susceptibility.
