## Supplementary File 2 for "Development of the Susceptibility-Spectrum Discrepancy Index (S2DI): A novel metric for antimicrobial stewardship in hospitalised patients"

**Supplementary information for**

**Development of the Susceptibility–Spectrum Discrepancy Index (S2DI): A novel metric for antimicrobial stewardship in hospitalised patients**

Shinya Tsuzuki, Ryuji Koizumi, Yusuke Asai, Yuuki Hashimoto, Norihiko Inoue, Yusuke Asai, Norio Ohmagari

**Figure S1. Daily transitions of the Susceptibility****–Spectrum Discrepancy Index (S2DI) scores and disposition outcomes during the first 14 days from the detection of bacteria among patients with *Escherichia coli* (left panel) and *Staphylococcus aureus* (right panel) bacteraemia**

Stacked bar charts show the distribution of S2DI scores on each hospital day. Coloured ribbons illustrate the day-to-day transitions in S2DI scores for individual patients together with disposition outcomes, including discharge (light grey), transfer/other (medium grey), and death (black).

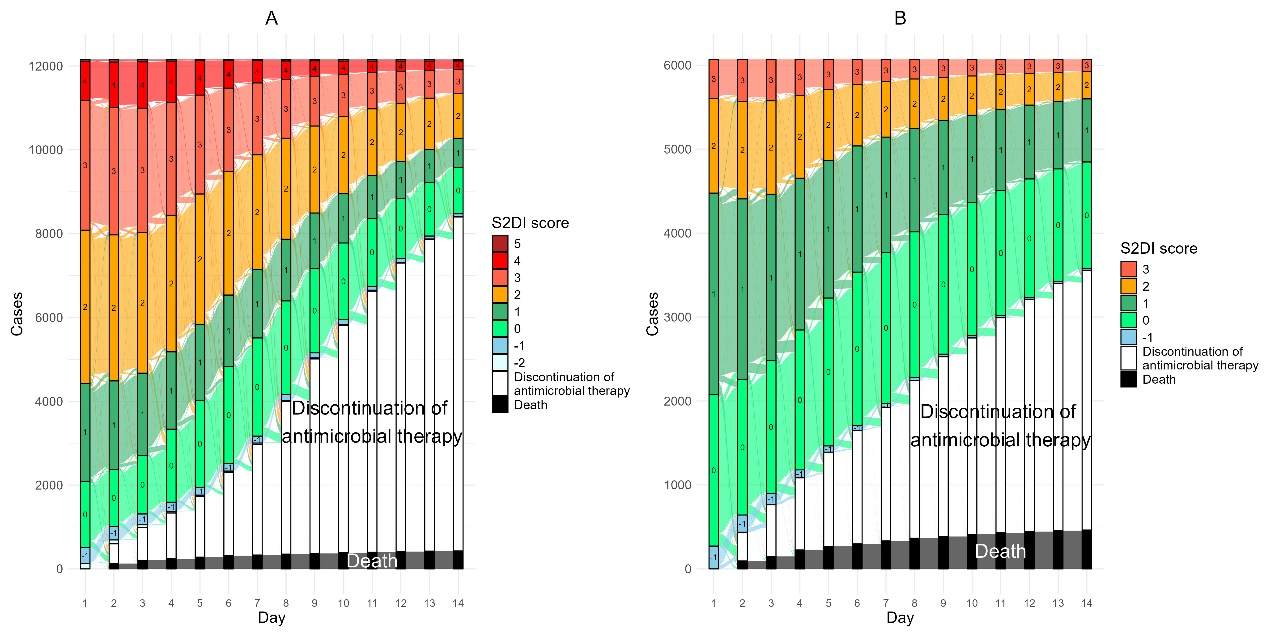

**Results of sensitivity analyses using data exclusively from facilities that provided a complete dataset for the 7-year period from 2017 to 2023.**

**Table S1-a. Characteristics of *Escherichia coli* bacteraemia patients extracted from the database (including cases that were discharged/died within 7 days of blood culture collection)**

| **Year** | **2017** | **2018** | **2019** | **2020** | **2021** | **2022** | **2023** | **Total** |
| --- | --- | --- | --- | --- | --- | --- | --- | --- |
| **Number of patients** | 1,236 | 1,419 | 1,701 | 1,827 | 1,965 | 2,013 | 2,069 | 12,230 |
| **Age [IQR] (min****–max)** | 79  [70-85]  (0-101) | 79  [70-86]  (0-101) | 79  [70-86]  (0-105) | 80  [72-86]  (0-107) | 80  [72-87]  (0-103) | 80  [72-87]  (0-101) | 80  [72-87]  (0-104) | 80  [72-86]  (0-107) |
| **Number of female patients (%)** | 673 (54.4) | 800 (56.3) | 948 (55.7) | 1,006 (55.1) | 1,074 (54.7) | 1,115 (55.4) | 1,148 (55.5) | 6,764  (55.3) |
| **Charlson comorbidity index [IQR]** **(min–max)** | 1  [0-2]  (0-11) | 1  [0-2]  (0-9) | 1  [0-2]  (0-9) | 1  [0-2]  (0-9) | 1  [0-2]  (0-9) | 1  [0-2]  (0-10) | 1  [0-2]  (0-9) | 1  [0-2]  (0-11) |
| **Clinical outcome, n (%)** |  |  |  |  |  |  |  |  |
| **Discharged alive** | 841 (68.0) | 968 (68.2) | 1,198 (70.4) | 1,308 (71.6) | 1,368 (69.6) | 1,368 (68.0) | 1,489 (72.0) | 8,540 (69.8) |
| **Transferred** | 225 (18.2) | 278 (19.6) | 315 (18.5) | 330 (18.1) | 402 (20.5) | 394 (19.6) | 375 (18.1) | 2,319 (19.0) |
| **Died** | 169 (13.6) | 172 (12.1) | 188 (11.1) | 188 (10.3) | 191 (9.8) | 244 (12.1) | 203 (9.8) | 1,355 (11.1) |
| **Unknown** | 1 (0.1) | 1 (0.1) | 0 (0) | 1 (0.1) | 4 (0.2) | 7 (0.3) | 2 (0.1) | 16 (0.1) |
| **30-day all-cause deaths, n (%)** | 79 (6.4) | 67 (4.7) | 90 (5.3) | 88 (4.8) | 100 (5.1) | 139 (6.9) | 126 (6.1) | 689 (5.6) |
| **S2DI score [IQR]** | 2  [0-3] | 2  [0-3] | 2  [1-3] | 2  [0-2] | 2  [0-3] | 2  [0-2] | 2  [0-2] | 2  [0-3] |
| **Days from admission to the detection of bacteria [IQR]** | 1  [1-8] | 1  [1-7] | 1  [1-5] | 1  [1-5] | 1  [1-3] | 1  [1-3] | 1  [1-2] | 1  [1-4] |
| **Length of stay* [IQR]** | 21  [13-42] | 19  [13-35] | 18  [12-35] | 18  [12-34] | 18  [11-34] | 17  [11-32] | 18  [12-32] | 18  [12-32] |

IQR: interquartile range; min: minimum; max: maximum; n: number; S2DI: Susceptibility**–**Spectrum Discrepancy Index

*Cases involving deaths and those currently hospitalised were excluded.

**Table S1-b. Characteristics of *Staphylococcus aureus* bacteraemia patients extracted from the database (including cases that were discharged/died within 7 days of blood culture collection)**

| **Year** | **2017** | **2018** | **2019** | **2020** | **2021** | **2022** | **2023** | **Total** |
| --- | --- | --- | --- | --- | --- | --- | --- | --- |
| **Number of patients** | 648 | 770 | 919 | 855 | 895 | 1,050 | 1,020 | 6,157 |
| **Age [IQR] (min****–max)** | 75  [65-84]  (0-97) | 74  [63-82]  (0-102) | 76  [63-84]  (0-103) | 76  [63-84]  (0-101) | 77  [67-85]  (0-101) | 79  [68-86]  (0-100) | 76  [66-85]  (0-98) | 76  [65-84]  (0-103) |
| **Number of female patients (%)** | 249 (38.4) | 274 (35.6) | 351 (38.2) | 302 (35.3) | 323 (36.1) | 424 (40.4) | 409 (40.1) | 2,332  (37.9) |
| **Charlson comorbidity index [IQR] (min–max)** | 1  [0-2]  (0-9) | 1  [0-2]  (0-9) | 1  [0-2]  (0-9) | 1  [0-2]  (0-10) | 1  [0-2]  (0-9) | 1  [0-2]  (0-10) | 1  [0-2]  (0-10) | 1  [0-2]  (0-10) |
| **Clinical outcome, n (%)** |  |  |  |  |  |  |  |  |
| **Discharged alive** | 294 (45.4) | 348 (45.2) | 392 (42.7) | 359 (42.0) | 380 (42.5) | 417 (39.7) | 452 (44.3) | 2,642 (42.9) |
| **Transferred** | 174 (26.9) | 212 (27.5) | 277 (30.1) | 261 (30.5) | 259 (28.9) | 300 (28.6) | 294 (28.8) | 1,777 (28.9) |
| **Died** | 180 (27.8) | 210 (27.3) | 250 (27.2) | 235 (27.5) | 255 (28.5) | 327 (31.1) | 263 (25.8) | 1,720 (27.9) |
| **Unknown** | 0 (0) | 0 (0) | 0 (0) | 0 (0) | 1 (0.1) | 6 (0.6) | 11 (1.1) | 18 (0.3) |
| **30-day all-cause deaths, n (%)** | 77 (11.9) | 90 (11.7) | 111 (12.1) | 127 (14.9) | 132 (14.7) | 175 (16.7) | 151 (14.8) | 863 (14.0) |
| **S2DI score [IQR]** | 1  [0-1] | 1  [0-1] | 1  [0-1] | 1  [0-1] | 1  [0-1] | 1  [0-1] | 1  [0-1] | 1  [0-1] |
| **Days from admission to the detection of bacteria [IQR]** | 5  [1-22] | 4  [1-22] | 4  [1-19] | 4  [1-20] | 3  [1-17.5] | 4  [1-20] | 3  [1-15] | 4  [1-19] |
| **Length of stay* [IQR]** | 43  [26-76] | 44  [24-79] | 42  [22-67] | 43  [23-75] | 38  [21-66] | 40  [23-68] | 37.5  [21-63] | 41  [23-69] |

IQR: interquartile range; min: minimum; max: maximum; n: number; S2DI: Susceptibility**–**Spectrum Discrepancy Index

*Cases involving deaths and those currently hospitalised were excluded.

**Table S2-a. Characteristics of *Escherichia coli* bacteraemia patients extracted from the facilities that provided complete datasets for 7 consecutive years**

| **Year** | **2017** | **2018** | **2019** | **2020** | **2021** | **2022** | **2023** | **Total** |
| --- | --- | --- | --- | --- | --- | --- | --- | --- |
| **Number of patients** | 938 | 1,075 | 1,122 | 1,183 | 1,301 | 1,275 | 1,309 | 8,203 |
| **Age [IQR] (min****–max)** | 79  [70-85]  (0-101) | 79  [71-86]  (0-101) | 79  [71-85]  (0-100) | 80  [72-86]  (0-107) | 80  [72-87]  (0-103) | 80  [72-87]  (0-101) | 80  [72-86]  (0-100) | 80  [72-86]  (0-107) |
| **Number of female patients (%)** | 497 (53.0) | 597 (55.5) | 637 (56.8) | 643 (54.4) | 706 (54.3) | 720 (56.5) | 729 (55.7) | 4,529  (55.2) |
| **Charlson comorbidity index [IQR] (min–max)** | 1  [0-2]  (0-11) | 1  [0-2]  (0-9) | 1  [0-2]  (0-9) | 1  [0-2]  (0-9) | 1  [0-2]  (0-9) | 1  [0-2]  (0-8) | 1  [0-2]  (0-9) | 1  [0-2]  (0-11) |
| **Clinical outcome, n (%)** |  |  |  |  |  |  |  |  |
| **Discharged alive** | 646 (68.9) | 748 (69.6) | 791 (70.5) | 860 (72.7) | 919 (70.6) | 907 (71.1) | 975 (74.5) | 5,846 (71.3) |
| **Transferred** | 179 (19.1) | 209 (19.4) | 232 (20.7) | 234 (19.8) | 278 (21.4) | 249 (19.5) | 242 (18.5) | 1,623 (19.8) |
| **Died** | 112 (11.9) | 117 (10.9) | 99 (8.8) | 89 (7.5) | 103 (7.9) | 111 (6.3) | 88 (6.7) | 719 (8.8) |
| **Unknown** | 1 (0.1) | 1 (0.1) | 0 (0) | 0 (0) | 1 (0.1) | 8 (0.6) | 4 (0.3) | 15 (0.2) |
| **30-day all-cause deaths, n (%)** | 39 (4.2) | 32 (3.0) | 38 (3.4) | 24 (2.0) | 44 (3.4) | 55 (4.3) | 48 (3.7) | 280 (3.4) |
| **S2DI score [IQR]** | 2  [0-3] | 2  [0-3] | 2  [1-3] | 2  [0-2] | 2  [0-3] | 2  [0-2] | 2  [0-2] | 2  [0-2] |
| **Days from admission to the detection of bacteria [IQR]** | 1  [1-6] | 1  [1-3] | 1  [1-2] | 1  [1-2] | 1  [1-2] | 1  [1-2] | 1  [1-1] | 1  [1-2] |
| **Length of stay* [IQR]** | 21  [15-40] | 19  [14-34] | 19  [13-34] | 19  [13-32] | 19  [13-32] | 18  [13-32] | 18  [13-31] | 19  [13-33] |

IQR: interquartile range; min: minimum; max: maximum; n: number; S2DI: Susceptibility**–**Spectrum Discrepancy Index

*Cases involving deaths and those currently hospitalised were excluded.

**Table S2-b. Characteristics of *Staphylococcus aureus* bacteraemia patients extracted from the facilities that provided complete datasets for 7 consecutive years**

| **Year** | **2017** | **2018** | **2019** | **2020** | **2021** | **2022** | **2023** | **Total** |
| --- | --- | --- | --- | --- | --- | --- | --- | --- |
| **Number of patients** | 489 | 486 | 575 | 503 | 510 | 602 | 574 | 3,739 |
| **Age [IQR] (min****–max)** | 75  [64-83]  (0-97) | 72  [63-83]  (0-102) | 76  [63-84]  (0-103) | 75  [62-84]  (0-101) | 77  [65-84]  (0-99) | 77  [67-85]  (0-100) | 75  [63-83]  (0-98) | 75  [64-84]  (0-103) |
| **Number of female patients (%)** | 187 (38.2) | 176 (36.2) | 225 (39.1) | 176 (35.0) | 183 (35.9) | 240 (39.9) | 206 (35.9) | 1,393  (37.3) |
| **Charlson comorbidity index [IQR] (min–max)** | 1  [0-2]  (0-9) | 1  [0-2]  (0-9) | 1  [0-2]  (0-9) | 1  [0-2]  (0-8) | 1  [0-2]  (0-9) | 1  [0-2]  (0-10) | 1  [0-2]  (0-9) | 1  [0-2]  (0-10) |
| **Clinical outcome, n (%)** |  |  |  |  |  |  |  |  |
| **Discharged alive** | 231 (47.2) | 242 (49.8) | 251 (43.7) | 222 (44.1) | 227 (44.5) | 264 (43.9) | 282 (49.1) | 1,719 (46.0) |
| **Transferred** | 135 (27.6) | 140 (28.8) | 186 (32.3) | 166 (33.0) | 156 (30.6) | 175 (29.1) | 182 (31.7) | 1,140 (30.5) |
| **Died** | 123 (25.2) | 104 (21.4) | 138 (24.0) | 115 (22.9) | 126 (24.7) | 160 (26.6) | 106 (18.5) | 872 (23.3) |
| **Unknown** | 0 (0) | 0 (0) | 0 (0) | 0 (0) | 1 (0.2) | 3 (0.5) | 4 (0.7) | 8 (0.2) |
| **30-day all-cause deaths, n (%)** | 46 (9.4) | 34 (7.0) | 42 (7.3) | 48 (9.5) | 60 (11.8) | 79 (13.1) | 49  (8.5) | 358 (9.6) |
| **S2DI score [IQR]** | 1  [0-1] | 1  [0-1] | 1  [0-1] | 1  [0-1] | 1  [0-1] | 1  [0-1] | 1  [0-1] | 1  [0-1] |
| **Days from admission to the detection of bacteria [IQR]** | 3  [1-19] | 2  [1-15] | 3  [1-17] | 3  [1-17] | 2  [1-15] | 2  [1-15] | 2  [1-11] | 2  [1-15] |
| **Length of stay* [IQR]** | 44  [29-76] | 43  [25-75] | 41  [23-65] | 41  [24-69] | 36  [21-60] | 39  [23-63] | 34  [21-62] | 40  [23-66] |

IQR: interquartile range; min: minimum; max: maximum; n: number; S2DI: Susceptibility**–**Spectrum Discrepancy Index

*Cases involving deaths and those currently hospitalised were excluded.

**Table S3-a. Characteristics of *Escherichia coli* bacteraemia patients by S2DI score (including cases that were discharged/died within 7 days from blood culture collection)**

| **Score** | **˗2** | **˗1** | **0** | **1** | **2** | **3** | **4** | **5** | **NA** | **Total** |
| --- | --- | --- | --- | --- | --- | --- | --- | --- | --- | --- |
| **Number of patients** | 37 | 170 | 2,412 | 1,715 | 2,833 | 1,815 | 544 | 37 | 2,667 | 12,230 |
| **Age [IQR] (min****–max)** | 77  [72-82]  (31-92) | 81  [73-86]  (27-97) | 81  [73-87]  (0-103) | 81  [73-87]  (0-107) | 80  [71-86]  (0-105) | 80  [71-86]  (0-102) | 78  [69.25-85]  (0-101) | 77  [67-84]  (49-96) | 79  [70-86] (0-105) | 80  [72-86]  (0-107) |
| **Number of female patients (%)** | 22  (59.5) | 86  (50.6) | 1434  (59.5) | 871  (50.8) | 1657  (48.8) | 886  (54.4) | 296  (37.8) | 14  (37.8) | 1492  (55.9) | 6758  (55.3) |
| **Charlson comorbidity index [IQR] (min–max)** | 1  [1-2]  (0-8) | 1  [0-2]  (0-9) | 1  [0-2]  (0-10) | 1  [0-2]  (0-11) | 1  [0-2]  (0-9) | 1  [0-2]  (0-9) | 1  [0-2]  (0-6) | 1  [0-2]  (0-6) | 1  [0-2]  (0-11) | 1  [0-2]  (0-11) |
| **Clinical outcome, n (%)** |  |  |  |  |  |  |  |  |  |  |
| **Discharged alive** | 24  (64.9) | 83  (48.8) | 1808  (75.0) | 1167  (68.0) | 2051  (72.4) | 1315  (72.5) | 373  (68.6) | 20  (54.1) | 1693  (63.5) | 8534  (69.8)) |
| **Transferred** | 7  (18.9) | 49  (28.8) | 440  (18.2) | 355  (20.7) | 562  (19.8) | 319  (17.6) | 106  (19.5) | 8  (21.6) | 467  (17.5) | 2313  (18.9) |
| **Died** | 6  (16.2) | 38  (22.4) | 160  (6.6) | 188  (11.0) | 212  (7.5) | 178  (9.8) | 63  (11.6) | 9  (24.3) | 501  (18.8) | 1355  (11.1) |
| **Unknown** | 0  (0.0) | 0  (0.0) | 4  (0.2) | 5  (0.3) | 8  (0.3) | 3  (0.2) | 2  (0.4) | 0  (0.0) | 6  (0.2) | 28  (0.2) |
| **30-day all-cause deaths, n (%)** | 0  (0.0) | 8  (4.7) | 28  (1.2) | 62  (3.6) | 86  (3.0) | 70  (3.9) | 29  (5.3) | 3  (8.1) | 1  (0.0) | 287  (2.3) |
| **Days from admission to the detection of bacteria [IQR]** | 1  [1-26] | 9.5  [1-33.75] | 1  [1-1] | 1  [1-4] | 1  [1-2] | 1  [1-2] | 1  [1-2] | 6  [1-23] | 1  [1-15] | 1  [1-4] |
| **Length of stay* [IQR]** | 28  [16-50] | 30  [16.75-63.5] | 17  [13-30.25] | 20  [14-36] | 19  [13-33] | 18  [12-33] | 25  [16-45] | 44  [24.5-69.25] | 12  [7-36] | 18  [12-34  ] |

IQR: interquartile range; min: minimum; max: maximum; n: number; S2DI: Susceptibility**–**Spectrum Discrepancy Index

*Cases involving deaths and those currently hospitalised were excluded.

**Table S3-b. Characteristics of *Staphylococcus aureus* bacteraemia patients by S2DI score (including cases that were discharged/died within 7 days from blood culture collection)**

| **Score** | **˗1** | **0** | **1** | **2** | **3** | **NA** | **Total** |
| --- | --- | --- | --- | --- | --- | --- | --- |
| **Number of patients** | 54 | 1,962 | 1,502 | 710 | 277 | 1,652 | 6,157 |
| **Age [IQR] (min****–max)** | 77.5  [66.5-86]  (30-100) | 75  [63-84]  (0-103) | 78  [67-85]  (0-101) | 76  [65-83]  (2-100) | 73  [63-82.25]  (0-97) | 78  [68-85] (0-102) | 76  [65-84]  (0-103) |
| **Number of female patients (%)** | 18  (33.3) | 755  (38.5) | 547  (36.4) | 257  (36.2) | 96  (34.7) | 659  (39.9) | 2,332  (37.9) |
| **Charlson comorbidity index [IQR] (min–max)** | 1  [0-2]  (0-9) | 1  [0-2]  (0-10) | 1  [0-2]  (0-10) | 1  [0-2]  (0-9) | 1  [0-2]  (0-7) | 1  [0-2]  (0-10) | 1  [0-2]  (0-10) |
| **Clinical outcome, n (%)** |  |  |  |  |  |  |  |
| **Discharged alive** | 20  (37.0) | 987  (50.3) | 648  (43.1) | 274  (38.6) | 113  (40.8) | 600  (36.3) | 2,642  (42.9) |
| **Transferred** | 15  (27.8) | 575  (29.3) | 457  (30.4) | 228  (32.1) | 88  (31.8) | 414  (25.1) | 1,777  (28.9) |
| **Died** | 19  (35.2) | 395  (20.1) | 394  (26.2) | 208  (29.3) | 75  (27.1) | 629  (38.1) | 1,720  (27.9) |
| **Unknown** | 0  (0.0) | 5  (0.3) | 3  (0.2) | 0  (0.0) | 1  (0.4) | 9  (0.5) | 18  (0.3) |
| **30-day all-cause deaths, n (%)** | 0  (0.0) | 152  (7.7) | 153  (10.2) | 86  (12.1) | 39  (14.1) | 433  (26.2) | 863  (14.0) |
| **Days from admission to the detection of bacteria [IQR]** | 38.5  [10.75-65.5] | 2  [1-13] | 3  [1-21] | 5  [1-22] | 1  [1-9] | 8  [1-23] | 4  [1-19] |
| **Length of stay* [IQR]** | 87  [44-148.5] | 40  [23-66] | 42  [23-69] | 43  [25.25-70.75] | 47  [28-76] | 38  [18-70.75] | 41  [23-69] |

IQR: interquartile range; min: minimum; max: maximum; n: number; S2DI: Susceptibility**–**Spectrum Discrepancy Index

*Cases involving deaths and those currently hospitalised were excluded.

**Table S4-a. Characteristics of *Escherichia coli* bacteraemia patients by S2DI score from the facilities that provided complete datasets for 7 consecutive years**

| **Score** | **˗2** | **˗1** | **0** | **1** | **2** | **3** | **4** | **5** | **Total** |
| --- | --- | --- | --- | --- | --- | --- | --- | --- | --- |
| **Number of patients** | 35 | 149 | 2,072 | 1,456 | 2,451 | 1,565 | 443 | 32 | 8,203 |
| **Age [IQR] (min****–max)** | 77  [72.5-82]  (31-92) | 81  [72-86]  (27-97) | 80  [73-86]  (0-103) | 80  [73-87]  (0-107) | 79  [71-86]  (0-105) | 79  [71-86]  (0-102) | 77  [69-85]  (0-101) | 77.5  [69-84.25]  (49-94) | 80  [72-86]  (0-107) |
| **Number of female patients (%)** | 22  (62.9) | 77  (51.7) | 1246  (60.1) | 732  (50.3) | 1438  (58.7) | 764  (48.8) | 238  (53.7) | 12  (37.5) | 4529  (55.2) |
| **Charlson comorbidity index [IQR] (min–max)** | 1  [0.5-2]  (0-8) | 1  [0-2]  (0-9) | 1  [0-2]  (0-9) | 1  [0-2]  (0-11) | 1  [0-2]  (0-8) | 1  [0-2]  (0-9) | 1  [0-2]  (0-9) | 1  [0-2]  (0-6) | 1  [0-2]  (0-11) |
| **Clinical outcome, n (%)** |  |  |  |  |  |  |  |  |  |
| **Discharged alive** | 23  (65.7) | 75  (50.3) | 1543  (74.5) | 991  (68.1) | 1767  (72.1) | 1129  (72.1) | 300  (67.7) | 18  (56.3) | 5846  (71.3) |
| **Transferred** | 7  (20.0) | 42  (28.2) | 399  (19.3) | 306  (21.0) | 491  (20.0) | 282  (18.0) | 91  (20.5) | 5  (15.6) | 1623  (19.8) |
| **Died** | 5  (14.3) | 32  (21.5) | 128  (6.2) | 155  (10.6) | 187  (7.6) | 152  (9.7) | 51  (11.5) | 9  (28.1) | 719  (8.8) |
| **Unknown** | 0  (0.0) | 0  (0.0) | 2  (0.1) | 4  (0.3) | 6  (0.2) | 2  (0.1) | 1  (0.2) | 0  (0.0) | 15  (0.2) |
| **30-day all-cause deaths, n (%)** | 0  (0.0) | 7  (4.7) | 52  (2.5) | 51  (3.5) | 79  (3.2) | 65  (4.2) | 23  (5.2) | 3  (9.4) | 280  (3.4) |
| **Days from admission to the detection of bacteria [IQR]** | 1  [1-26.5] | 7  [1-30] | 1  [1-1] | 1  [1-4] | 1  [1-2] | 1  [1-2] | 1  [1-3] | 5.5  [1-20.75] | 1  [1-2] |
| **Length of stay* [IQR]** | 28.5  [15.5-50.5] | 29  [16-53] | 17  [13-30] | 19  [14-34] | 19  [13-32] | 18  [12-33] | 25  [16-43] | 40  [24-67] | 19  [13-33] |

IQR: interquartile range; min: minimum; max: maximum; n: number; S2DI: Susceptibility**–**Spectrum Discrepancy Index

*Cases involving deaths and those currently hospitalised were excluded.

**Table S4-b. Characteristics of *Staphylococcus aureus* bacteraemia patients by S2DI score from the facilities that provided complete datasets for 7 consecutive years**

| **Score** | **˗1** | **0** | **1** | **2** | **3** | **Total** |
| --- | --- | --- | --- | --- | --- | --- |
| **Number of patients** | 44 | 1,655 | 1,228 | 590 | 222 | 3,739 |
| **Age [IQR] (min****–max)** | 76.5  [64.5-83]  (30-93) | 75  [62.75-83]  (0-103) | 77  [66-85]  (0-101) | 75  [63-83]  (2-100) | 73  [63-82]  (0-96) | 75  [64-84]  (0-103) |
| **Number of female patients (%)** | 14  (31.8) | 634  (38.3) | 454  (37.0) | 215  (36.4) | 76  (34.2) | 1,393  (37.3) |
| **Charlson comorbidity index [IQR] (min–max)** | 1  [0-2]  (0-7) | 1  [0-2]  (0-9) | 1  [0-2]  (0-10) | 1  [0-2]  (0-10) | 1  [0-2]  (0-9) | 1  [0-2]  (0-10) |
| **Clinical outcome, n (%)** |  |  |  |  |  |  |
| **Discharged alive** | 18  (40.9) | 835  (50.5) | 533  (43.4) | 242  (41.0) | 91  (41.0) | 1,719  (46.0) |
| **Transferred** | 12  (27.3) | 485  (29.3) | 381  (31.3) | 189  (32.0) | 73  (32.9) | 1140  (30.5) |
| **Died** | 14  (31.8) | 331  (20.0) | 311  (25.3) | 159  (26.9) | 57  (25.7) | 872  (23.3) |
| **Unknown** | 0  (0.0) | 4  (0.2) | 3  (0.2) | 0  (0.0) | 1  (0.5) | 8  (0.2) |
| **30-day all-cause deaths, n (%)** | 0  (0.0) | 134  (8.1) | 128  (10.4) | 68  (11.5) | 28  (12.6) | 358  (9.6 ) |
| **Days from admission to the detection of bacteria [IQR]** | 33.5  [6.75-62] | 2  [1-12] | 2  [1-18] | 4  [1-20] | 1  [1-9] | 2  [1-15] |
| **Length of stay* [IQR]** | 80  [40.5-140] | 39.5  [23-65.25] | 39  [23-65.75] | 43  [24-65] | 43.5  [28-73.5] | 40  [23-66] |

IQR: interquartile range; min: minimum; max: maximum; n: number; S2DI: Susceptibility**–**Spectrum Discrepancy Index

*Cases involving deaths and those currently hospitalised were excluded.

**Results of sensitivity analyses using a random effects model on data from facilities that provided complete datasets for 7 consecutive years**

**Figure S2. Forest plots of fixed-effect estimates from mixed-effects logistic regression models for *Escherichia* *coli* (left panel) and *Staphylococcus aureus* (right panel) bacteraemia**

Each point represents the adjusted fixed-effect estimate on the log-odds scale, with horizontal bars showing the corresponding 95% confidence intervals. Covariates include patient-level factors (age, sex, Charlson comorbidity index), specific departments at the facilities, hospital bed size, geographic region (reference: Kanto, in which Tokyo is located), and calendar year. Positive estimates indicate a relative tendency toward broad-spectrum antibiotic use, whereas negative estimates indicate a tendency toward narrow-spectrum therapy. Estimates crossing zero suggest no statistically significant association.

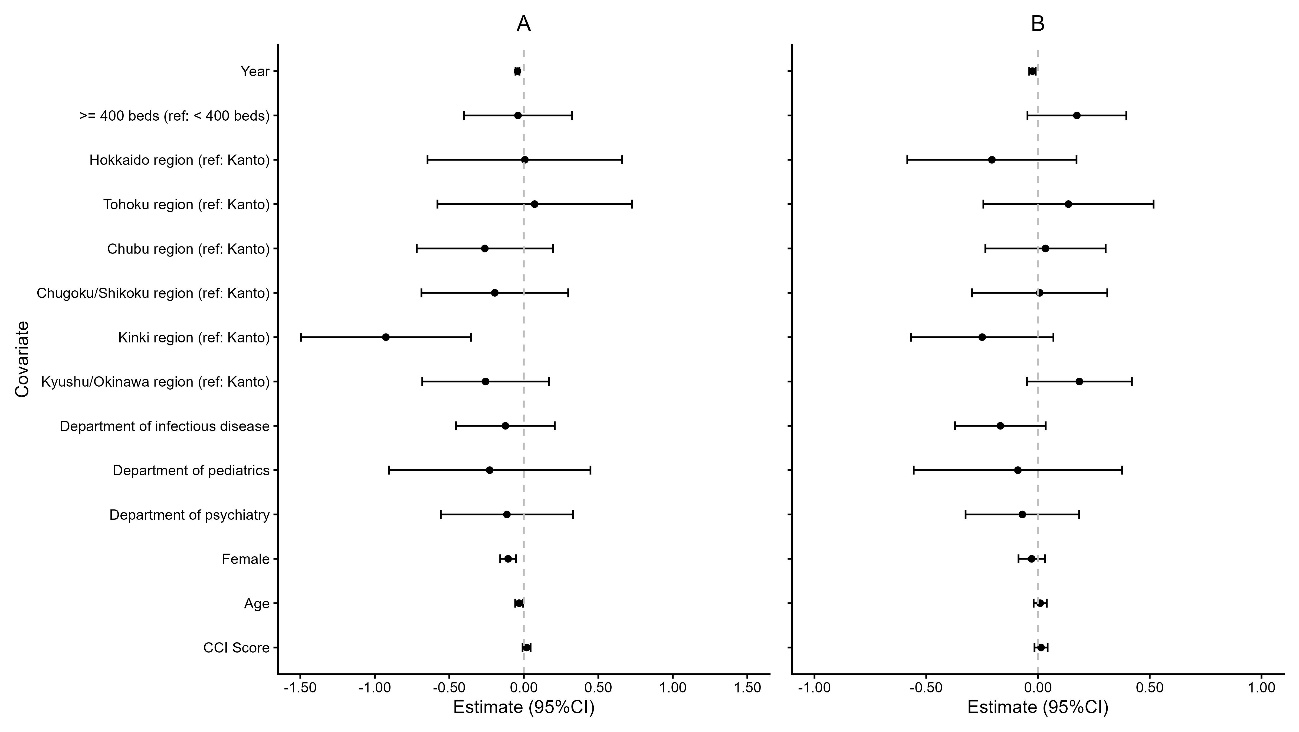

**Figure S3. Caterpillar plots of facility-level random intercepts from the mixed-effects models for *Escherichia coli* (left panel) and *Staphylococcus aureus* (right panel) bacteraemia**

Each point represents the best linear unbiased prediction (BLUP) of the facility-specific random intercept, with horizontal bars indicating the corresponding 95% confidence intervals. The random intercepts are centred at zero, which represents the average facility. Positive values indicate a greater tendency toward broad-spectrum antibiotic use, whereas negative values indicate a tendency toward narrow-spectrum use. Facilities are ordered by the magnitude of their BLUPs.

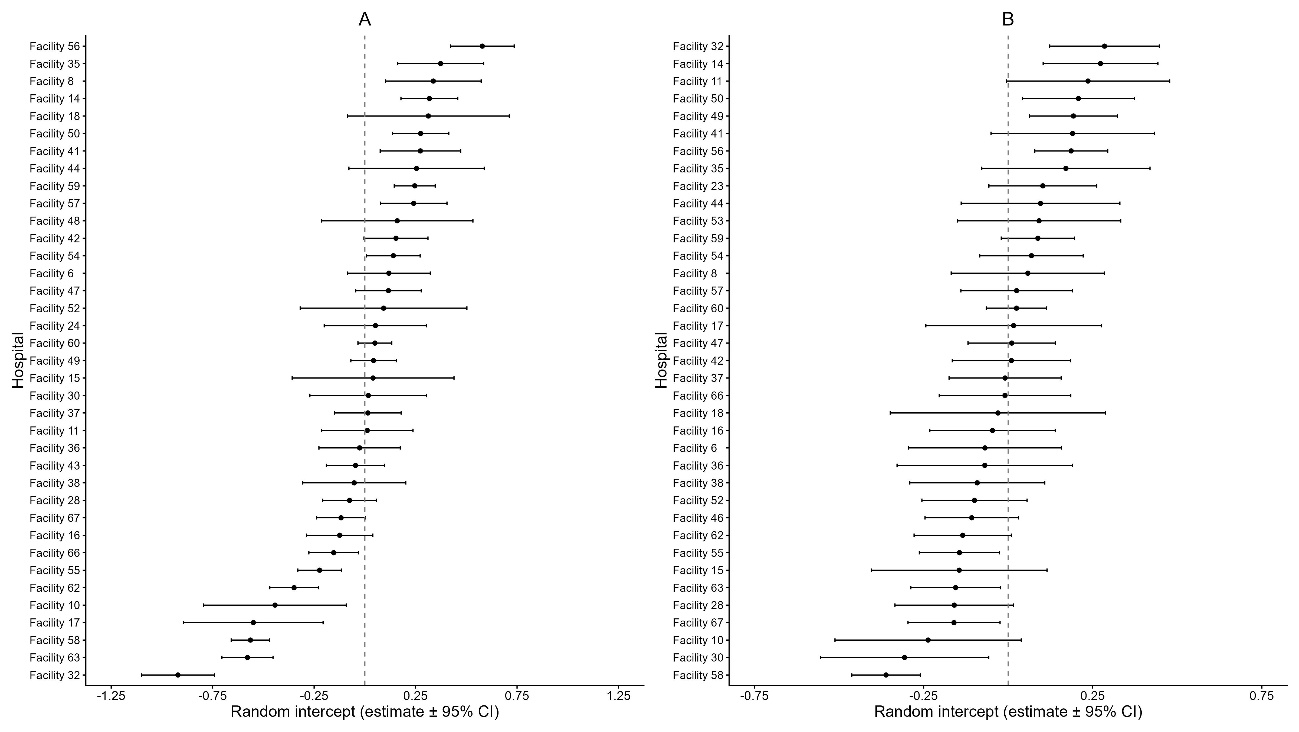

**Figure S4. Snail plots of facility-level random effects for broad- and narrow-spectrum antibiotic use in *Escherichia coli* (left panel) and *Staphylococcus aureus* (right panel) bacteraemia**

Each bar represents the best linear unbiased prediction (BLUP) of the facility-level random intercept from the mixed-effects model. Bars extending upward indicate a tendency toward broad-spectrum antibiotic use, whereas downward bars indicate a tendency toward narrow-spectrum antimicrobials. The magnitude of each bar reflects the relative contribution of each facility to between-facility variability. Dark bars denote broad-spectrum-leaning facilities and light bars denote narrow-spectrum-leaning facilities. Facility identifiers are shown around the circumference.

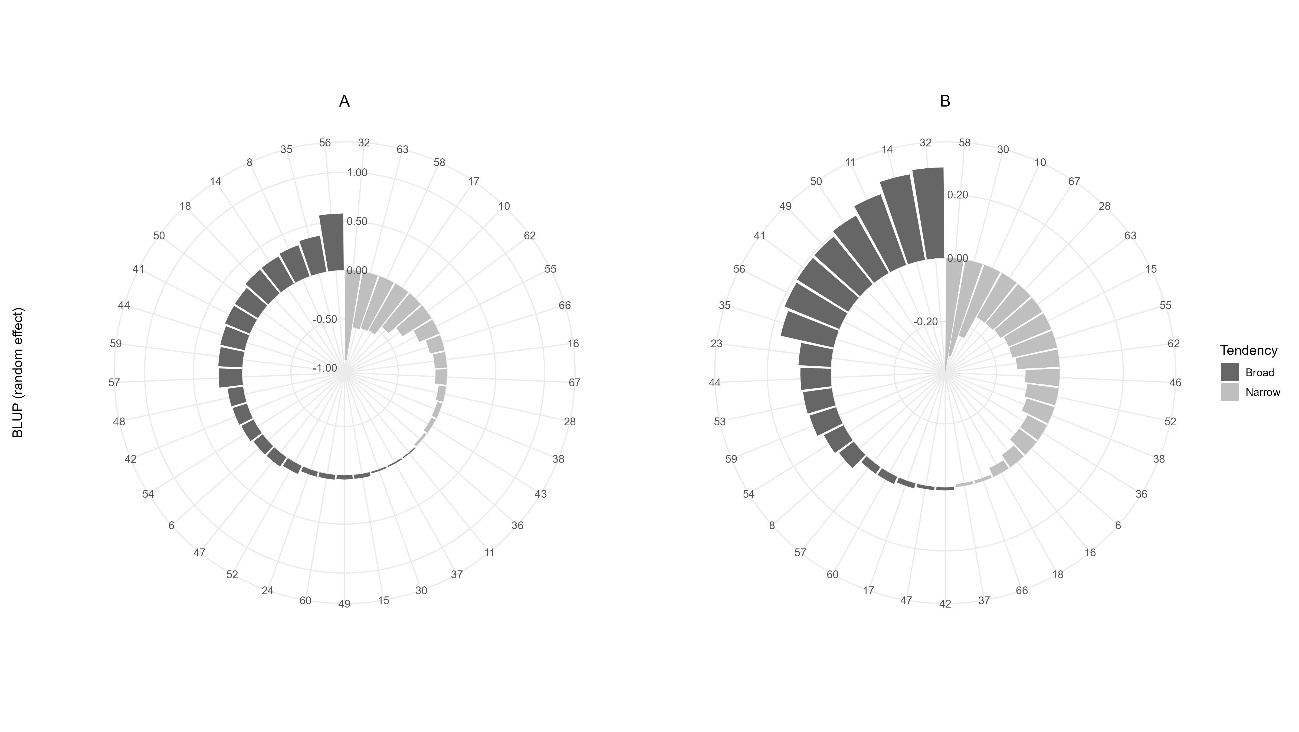
